# Multicenter validation of an assay to predict anti-PD-1 disease control in patients with recurrent or metastatic Head and Neck Squamous Cell Carcinoma: The PREDAPT Study

**DOI:** 10.1101/2024.05.31.24308285

**Authors:** Kevin C. Flanagan, Jon Earls, Jeffrey Hiken, Rachel L. Wellinghoff, Michelle M. Ponder, Howard L. Mcleod, William H. Westra, Vera Vavinskaya, Leisa Sutton, Ida Deichaite, Orlan K. Macdonald, Karim Welaya, James L. Wade, Georges Azzi, Andrew W. Pippas, Jennifer Slim, Bruce Bank, Xingwei Sui, Steven E. Kossman, Todd D. Shenkenberg, Warren L. Alexander, Katharine A Price, Jessica Ley, David N. Messina, Jarret I. Glasscock, A. Dimitrios Colevas, Ezra E.W. Cohen, Douglas R. Adkins, Eric J. Duncavage

## Abstract

**Background:** Despite advances in cancer care and detection, more than 65% of patients with squamous cell cancer of the head and neck (HNSCC) will develop recurrent and/or metastatic disease. The prognosis for these patients is poor with a 5 year overall survival of 39%. Recent treatment advances in immunotherapy, including immune checkpoint inhibitors like pembrolizumab and nivolumab, have resulted in clinical benefit in a subset of patients. There is a critical clinical need to identify patients who benefit from these anti-PD-1 immune checkpoint inhibitors.

**Methods:** Here we report findings from a multi-center observational study, PREDAPT (ClinicalTrials.gov: NCT04510129), conducted across 17 US healthcare systems. PREDAPT aimed to validate OncoPrism-HNSCC, a clinical biomarker assay predictive of disease control in recurrent or metastatic HNSCC patients treated with anti-PD-1 immune checkpoint inhibitors as a single agent (monotherapy) and in combination with chemotherapy (chemo-immunotherapy). The test used RNA-sequencing data and machine learning models to score each patient and place them into groups of Low, Medium, or High.

**Results:** The OncoPrism-HNSCC prediction significantly correlated with disease control in both the monotherapy cohort (n=62, p=0.004) and the chemo-immunotherapy cohort (n=50, p=0.01). OncoPrism-HNSCC also significantly predicted progression-free survival in both cohorts (p=0.015 and p=0.037, respectively). OncoPrism-HNSCC had more than threefold higher specificity than PD-L1 combined positive score and nearly fourfold higher sensitivity than tumor mutational burden for predicting disease control.

**Conclusions:** Here we demonstrate the clinical validity of the OncoPrism-HNSCC assay in identifying patients with disease control in response to anti-PD-1 immune checkpoint inhibitors.

**WHAT IS ALREADY KNOWN ON THIS TOPIC:** Anti-PD-1 immune checkpoint inhibitors such as pembrolizumab benefit a subset of patients with recurrent or metastatic head and neck squamous cell carcinoma (RM- HNSCC), but current biomarkers are inadequate at identifying these patients.

**WHAT THIS STUDY ADDS:** This study describes the validation of a new RNA-based test that predicts disease control and progression-free survival in response to anti-PD-1 therapy with high sensitivity and specificity. The test was validated using two independent cohorts of patients from 17 community and academic sites.

**HOW THIS STUDY MIGHT AFFECT RESEARCH, PRACTICE OR POLICY:** The test had significantly higher sensitivity than TMB and significantly higher specificity than PD-L1, enabling clinicians to make more informed decisions when prioritizing treatment. Use of the test has the potential to avoid unnecessary chemotherapy and/or anti-PD-1 treatment and improve patient outcomes.

## BACKGROUND

Head and neck squamous cell carcinomas (HNSCC) represent a significant healthcare burden. Worldwide, HNSCC is the seventh most common cancer with 870,000 new cases and 440,000 deaths[1]. More than 65% of these HNSCC patients are ultimately diagnosed with recurrent or metastatic disease[2,3]. Patients with recurrent or metastatic head and neck squamous cell carcinoma (RM-HNSCC) have a poor prognosis with median overall survival of just 10.7-13.0 months[4]. The introduction of anti-PD-1 immune checkpoint inhibitors (ICIs) such as pembrolizumab and nivolumab has improved outcomes for a subset of patients, but ICIs are associated with serious adverse reactions and a high financial burden to the health system[5–9]. In practice, many patients receive ICI in combination with platinum or other chemotherapies and choosing between ICI monotherapy and ICI in combination with chemotherapy (chemo- immunotherapy) is an important treatment decision.

Currently, anti-PD-1 treatments are recommended for patients with PD-L1 immunohistochemistry (IHC) combined positive score (CPS) of at least one. However, while PD-L1 IHC testing is recommended for all patients, PD-L1 status does not correlate well with clinical benefit, as measured by Disease Control Rate (DCR) defined by the fraction of patients without disease progression as best tumor response post-treatment. In KEYNOTE-048, for example, patients with PD-L1 CPS≥1 had only a 3% higher DCR from immunotherapy as a single agent and a 1% lower DCR from chemo- immunotherapy when compared to the same treatment arm regardless of PD-L1 CPS[4,10]. While Tumor Mutational Burden (TMB) is sometimes used to aid treatment decisions, its clinical utility in HNSCC is less clear[11–13]. ICI undoubtedly provides clinical benefit to a subset of patients, but current methods for identifying exactly which patients benefit are insufficient.

There is an unmet clinical need for more robust methods of predicting disease control in response to PD-1 inhibitors. Previously we described the development of an RNA-sequencing-based classifier to predict disease control with increased sensitivity and specificity compared to PD-L1 CPS in patients with RM-HNSCC treated with anti-PD-1 monotherapy[14]. In that study, we classified patients as progressors or non-progressors based on the median predicted probability of disease control as determined by the test. Building on that work, we refined the test and its thresholds for disease control prediction to create three groups correlated with predicted likelihood of disease control: Low, Medium, and High. Here we report the validation and test performance in two independent cohorts of patients, one treated with anti-PD-1 alone (monotherapy) and one with anti-PD-1 in combination with platinum-based chemotherapy (chemo- immunotherapy). The resulting laboratory developed test (LDT), OncoPrism-HNSCC, classifies patients into three groups, Low (25% of patients with lowest likelihood of disease control), Medium (25% of patients with indeterminate likelihood of disease control), and High (50% of patients with high likelihood of disease control). The test predicts disease control in response to anti-PD-1 treatments with both high sensitivity and specificity.

## METHODS

### Study Design and Participants

Patients were recruited from the following academic and community study sites across the United States, with the aim of a representative sample of the affected population: Washington University in St. Louis (St. Louis, MO), University of California San Diego (San Diego, CA), Intermountain Healthcare (Salt Lake City, UT), Gundersen Medical Foundation (La Crosse, WI), Cancer Care Northwest (Spokane, WA), Cox Medical Centers (Springfield, MO), Decatur Memorial Hospital (Decatur, IL), Holy Cross Hospital (Fort Lauderdale, FL), John B Amos Cancer Center (Columbus, GA), MultiCare Institute for Research and Innovation (Tacoma, WA), Northwest Oncology and Hematology (Hoffman Estates, IL), Ochsner Lafayette General Medical Center (Lafayette, LA), Providence Regional Cancer System (Lacey, WA), Sharp Clinical Oncology Research (San Diego, CA), Stanford University (Stanford, CA), William Beaumont Army Medical Center (Fort Bliss, TX), Baylor College of Medicine (Houston, TX), Brooke Army Medical Center (Fort Sam Houston, TX), Dayton Physicians Network (Dayton, OH), Mayo Clinic (Rochester, MN), Revive Research Institute (Sterling Heights, MI), and Valley Cancer Associates (Harlingen, TX).

Patients were enrolled from 2019-2023 in a combined prospective and retrospective, observational study following the inclusion and exclusion criteria outlined below. No patient level study data were reported to patients or physicians and the patients and public were not involved in the study design. Eligible patients had recurrent or metastatic histologically or cytologically confirmed HNSCC and were treated with curative intent with anti-PD-1 either as a single agent (monotherapy) or in combination with chemotherapy (chemo-immunotherapy) for recurrent or metastatic disease. Tissue specimens analyzed in the study were collected from pre-treatment tumor samples originally processed as formalin-fixed and paraffin embedded (FFPE) specimens using standard histologic protocols. De-identified, pre-treatment FFPE tumor biopsy specimens were provided to Cofactor Genomics for OncoPrism-HNSCC and PD-L1 IHC analysis. Following treatment, each patient’s tumor response to immunotherapy was evaluated using RECIST, PERCIST, or other clinical criteria as appropriate in standard of care to determine disease control. Patients with insufficient tissue for analysis (<10% tumor cells as determined by a study pathologist [EJD]) and samples with greater than 22.4 months between biopsy and treatment were excluded from the study[14]. Primary or metastatic tumor specimens were accepted, but metastatic tumors from liver or bone were not included due to confounding tissue RNA expression and the difficulty of recovering and processing decalcified FFPE RNA. Length of follow-up ranged from 34 days to 64 months. The study protocol, “A Multicenter Cancer Biospecimen Collection Study” is registered as “NCT04510129—PREDicting immunotherapy efficacy from Analysis of Pre-treatment Tumor biopsies (PREDAPT)” on clinicaltrials.gov. The study protocol was approved by institutional review boards at either the study (Advarra, Inc. [Columbia, MD] or WCG IRB [Puyallup, Washington]) or site level, as appropriate. All patients provided signed, informed consent to participate, or consent was waived for deceased patients according to study protocol. Independent data monitoring was conducted by the study clinical research organization Curebase, Inc. (San Francisco, CA).

### RNA Extraction

RNA was extracted using RNAstorm (Biotium, Fremont, CA) according to the manufacturer’s instructions. RNA quantity was assessed by the High Sensitivity RNA Qubit assay (Thermo Fisher Scientific, Waltham, MA). A predefined yield of 40 ng FFPE RNA was used as the minimum QC threshold. Quality of the RNA was assessed using a bioanalyzer (Agilent Technologies, Santa Clara, CA), and a DV200 of greater than 24% was required for all samples.

### Library Preparation and Sequencing

Libraries were prepared using the QuantSeq 3’ mRNA-Seq Library Prep Kit FWD for Illumina (Lexogen, Inc., Greenland, NH), following the manufacturer’s instructions.

Library RNA input was 40 ng for all samples. UMI Second Strand Synthesis Module for QuantSeq FWD (Lexogen, Inc., Greenland, NH) replaced Second Strand Synthesis Mix 1 in the workflow. All samples were processed with two OncoPrism-HNSCC positive controls and a No Template Control. The positive (high or medium scoring) controls were RNA extracted from RM-HNSCC samples as described above. Final libraries were sequenced to a minimum depth of 10 million single-end 75 base pair reads on a NextSeq500 (Illumina, San Diego, CA), following the manufacturer’s protocols.

### Immunohistochemistry

PD-L1 staining was performed by Mosaic Labs (Lake Forest, CA) using the 22C3 pharmDx antibody (Agilent Technologies, Inc., Santa Clara, CA) or NeoGenomics Laboratories (Fort Myers, FL) using the PD-L1 22C3 FDA (KEYTRUDA®) for HNSCC Head and Neck stain. CPS assessment was performed by WHW or by NeoGenomics. H&E staining was performed by NeoGenomics as part of the PD-L1 22C3 test or at Cofactor Genomics using xylene substitute Slide Brite (Newcomer Supply, Middleton, WI), as detailed by manufacturers.

### Processing of RNA sequencing data

FASTQ files were preprocessed with trim_galore/cutadapt version 0.4.1 to remove adapter sequences as well as reads with PHRED quality scores less than 20 and reads that were less than 20 basepairs. The trimmed reads were aligned to the human genome GRCh38 with STAR version 2.5.2a using the two-pass method as previously described[15]. Read counts were generated using htseq-count version 0.9.1 and annotation from Gencode version 22[15]. The data was normalized as counts per million (CPM) and log2 transformed using unique reads aligning to protein coding regions.

Samples were required to have a minimum of 30% exonic alignment and 800,000 unique deduplicated counts to be included in the study.

### Tumor Mutational Burden

Tumor Mutational Burden (TMB) was measured using the GatewaySeq (Washington University, St. Louis, MO) targeted DNA assay which was run in a CLIA-accredited clinical laboratory. GatewaySeq calculates TMB using the Illumina (San Diego, CA) Dragen TMB caller in tumor-only mode and non-synonymous TMB output. We used the clinically validated GatewaySeq definition of TMB High (20 or more mutations per megabase) to categorize patients as TMB High or TMB Low. Fifty to 250 ng DNA was used as input. DNA was extracted using DNAstorm (Biotium, Fremont, CA) according to the manufacturer’s instructions.

### Model Training

Data from 1205 total samples were used to select features, refine the protocol, train the model, and validate the model (Figure 1). Data from 790 patient samples were used to identify 149 candidate features related to immune response with detectable expression across two publicly available datasets (Table S2)[16,17]. An additional 415 patient samples were collected for the PREDAPT trial. Samples that were excluded or failed QC requirements are detailed in Table S1. The remaining PREDAPT samples (n=211) were divided into a training cohort and two validation cohorts based on treatment and time enrollment was completed. The training cohort consisted of 99 samples from patients receiving anti-PD-1 monotherapy at eleven PREDAPT healthcare systems[14]. A supervised machine learning, logistic regression model was built using this training dataset. Patients with complete response, partial response or stable disease were treated as the positive class. Samples from 34 patients ultimately assigned to validation Cohort 1 were used as a preliminary evaluation of the training model performance.

**Figure 1.**
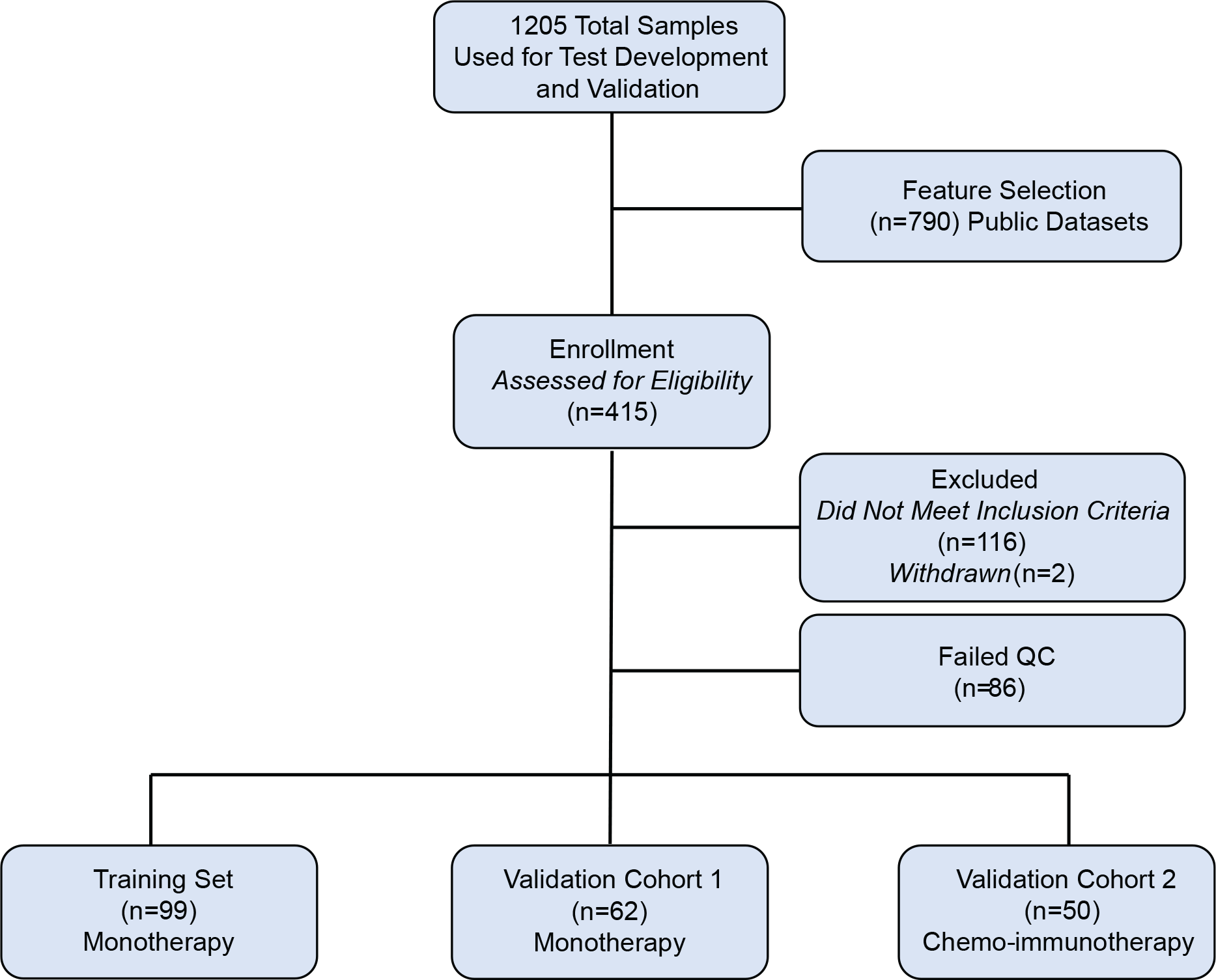
Samples used to develop, train, and validate OncoPrism-HNSCC. Data from a total of 1205 samples were used to select features, refine the protocol, train the model, and validate the model. Data from 790 publicly available samples were used to select features. 415 patient samples were collected as part of the PREDAPT trial. 116 samples were ineligible and were excluded from this study. 86 samples failed quality control and were excluded. Two patients were withdrawn from the study. Of the remaining patient samples, 161 were treated with monotherapy anti-PD-1 and 50 were treated with chemo-immunotherapy anti-PD-1. Of the monotherapy samples, 99 were used to train the model, and the remaining 62 monotherapy samples served as validation Cohort 1. The 50 chemo-immunotherapy samples served as validation Cohort 2.

### OncoPrism Scores and Prediction

The OncoPrism-HNSCC biomarker generates an OncoPrism Score from 0 to 100 that correlates with predicted disease control in patients with RM-HNSCC treated with anti- PD-1 monotherapy. Higher OncoPrism Scores represent higher confidence by the model that the patient will have disease control. The thresholds for the OncoPrism Groups were defined from the training data. Considering *n* unique patient samples, patients are chosen *n* times with replacement and used for training a model. Using this trained model, a score ("out-of-bag score") is generated for the remainder of patients[14,18]. This process was repeated 1000 times, and the out-of-bag score of each patient was averaged to generate a mean training OncoPrism Score. The threshold between the Low Group (OncoPrism Scores 0-37) and the Medium Group (OncoPrism Scores 38-52) is defined as the value of the 25th percentile mean score. The threshold between the Medium Group and the High Group (OncoPrism Scores 52- 100) is defined as the value of the 50th percentile mean score. These training cohort mean score thresholds are used for all subsequent validation and analysis to define the OncoPrism Groups.

### Validation of Performance

Clinical validation of the OncoPrism-HNSCC assay was performed using a separate cohort of 112 unique patient samples divided into two independent cohorts (Cohort 1 and Cohort 2). Samples were processed in the Cofactor Genomics CAP-accredited, CLIA-certified laboratory using strict quality controls. The primary validation metric was DCR in each OncoPrism Group. DCR was calculated by dividing the sum of patients with RECIST 1.1-defined categories of stable disease, partial response, and complete response as best response by the total number of patients in each group. DCR was used because of similar PFS and clinical benefit previously observed among patients with best response of stable disease and partial response[14]. To measure the test’s ability to enrich for disease control in response to anti-PD-1 monotherapy, 62 FFPE tumor samples from 62 monotherapy treated patients from 15 clinical sites were processed through the OncoPrism-HNSCC workflow (Cohort 1). As an additional independent validation, 50 FFPE tumor samples from 50 chemo-immunotherapy treated patients at 11 clinical sites were processed through the OncoPrism-HNSCC workflow (Cohort 2). Patient specimens came from 17 unique clinical sites in total. OncoPrism Scores were generated for each sample, and were assigned to the Low, Medium, or High OncoPrism Groups based on these scores. Operators were blinded to the RECIST label when processing samples and generating OncoPrism Scores. The RECIST labels for each patient were determined independently from the OncoPrism Group and were then used to determine the DCR for each group in the validation set.

For the purposes of test validation and treatment recommendations, Low Group patients are classified as predicted progressors, Medium Group patients are considered an indeterminate result, and High Group patients are classified as predicted to have disease control in response to ICI. The Medium Group is considered an indeterminate result due to the variation seen in Medium Group DCR and PFS across datasets (data not shown and Fig 2B-C & E-F). When comparing performance with PD-L1 CPS, the High Group patients are considered the predicted positive class (predicted disease control), while the Low and Medium Group patients are treated as the predicted negative class (no predicted disease control). Including the entire set of patients in these calculations allows for a direct comparison to PD-L1 CPS even though the intended use of OncoPrism-HNSCC is to consider a Medium Group result as indeterminate.

**Figure 2.**
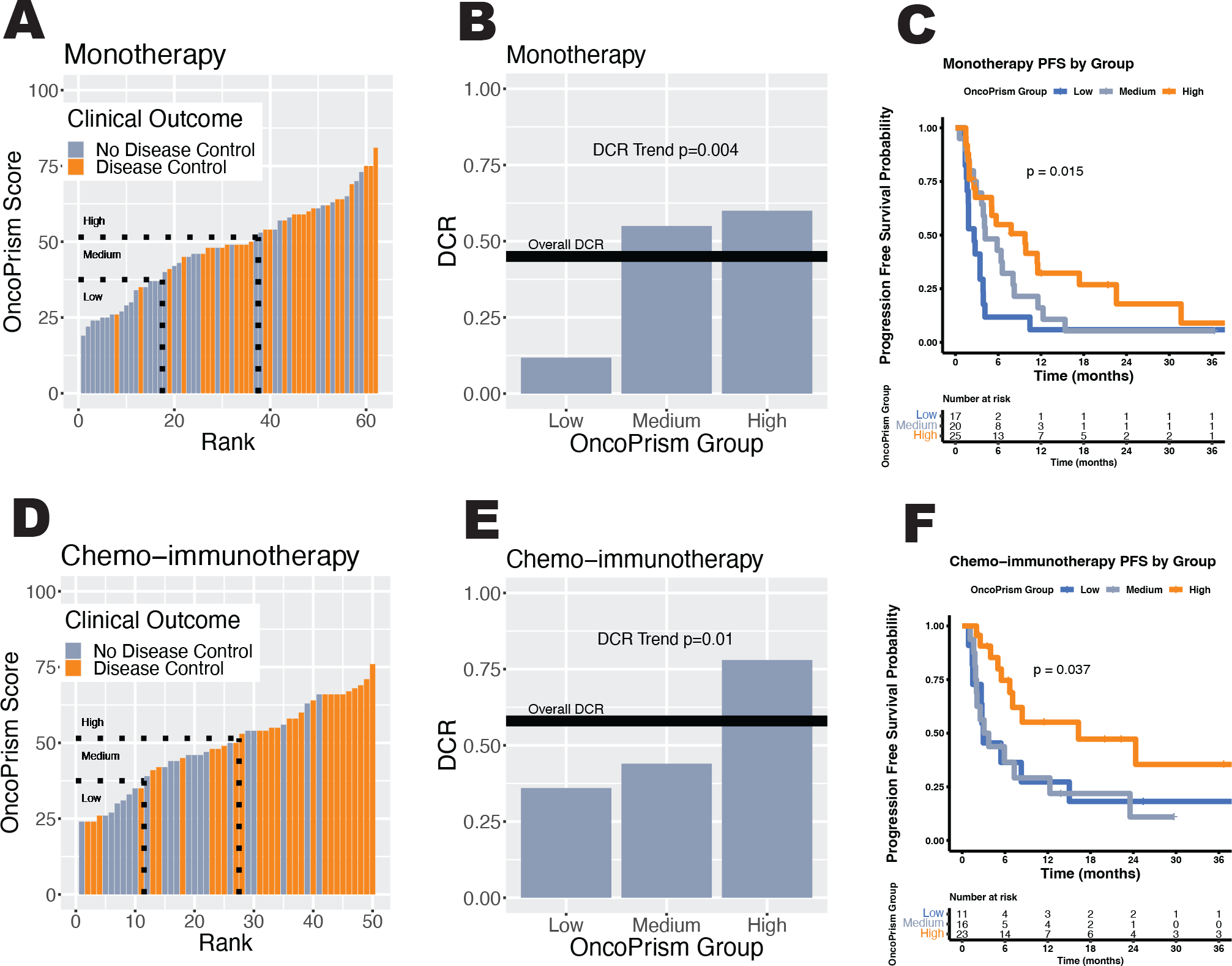
OncoPrism-HNSCC Score and Group are correlated with disease control in independent monotherapy and chemo-immunotherapy validation cohorts. Samples are ordered by their OncoPrism Score for the monotherapy-treated **(A)** and chemo- immunotherapy **(D)** validation cohorts. Lower scores are more likely to be progressors (grey) while higher scores are more likely to have disease control (orange). Based on their OncoPrism Score and predetermined thresholds (dotted lines), each patient sample is assigned to an OncoPrism Group (Low, Medium, or High). OncoPrism Groups are significantly correlated with disease control rate (DCR) in the monotherapy (p=0.004) **(B)** and chemo-immunotherapy (p=0.01) **(E)** validation cohorts. P-values for the significance of the trend were calculated using Cochrane-Armitage. OncoPrism Groups are significantly correlated with Progression-Free Survival (PFS) in the monotherapy (p=0.015) **(C)** and chemo-immunotherapy (p=0.037) **(F)** validation cohorts. P-values for PFS were calculated using log rank methods.

### Statistics

The primary endpoint of this study was disease control rate. A two-sided Cochran-Armitage test for trends was used to test the significance of the trend of increasing proportions for the DCRs of OncoPrism Groups. Power analysis was performed using the training cohort out-of-bag area under the ROC curve, seeking a two-sided Type 1 error (alpha) of 0.05 and a Type 2 error (beta) of 0.80. A minimum sample size of 36 was calculated to power the primary endpoint (Cohran-Armitage Test for trend in proportions for DCR). Expecting that training cohort out-of-bag performance may overestimate independent cohort performance, we sought a minimum of 50 samples for each cohort. PFS was defined as the time from start of ICI treatment to progression or death. Patients were censored if they had not progressed at last follow up. One OncoPrism High Group patient treated with chemo-immunotherapy was excluded because of an unknown date of progression. PFS figures and analysis were done using the “survminer” and “survival” packages, and significance was determined using log rank methods [19,20]. Differences in sensitivity and specificity were tested using McNemar’s test. 95% Confidence intervals for model performance metrics were calculated using a non-parametric bootstrap resampling method. In all cases a p-value of less than 0.05 was considered significant.

## RESULTS

Patients meeting all inclusion criteria (n=211) were divided into a training cohort (n=99) and two validation cohorts (Cohort 1, n=62 and Cohort 2, n=50; Figure 1; Table S3).

The training and validation cohorts have similar patient and disease characteristics, except that the training cohort and validation Cohort 1 were treated with monotherapy ICI while validation Cohort 2 was treated with ICI in combination with chemotherapy (chemo-immunotherapy; Table 1).

**Table 1:**
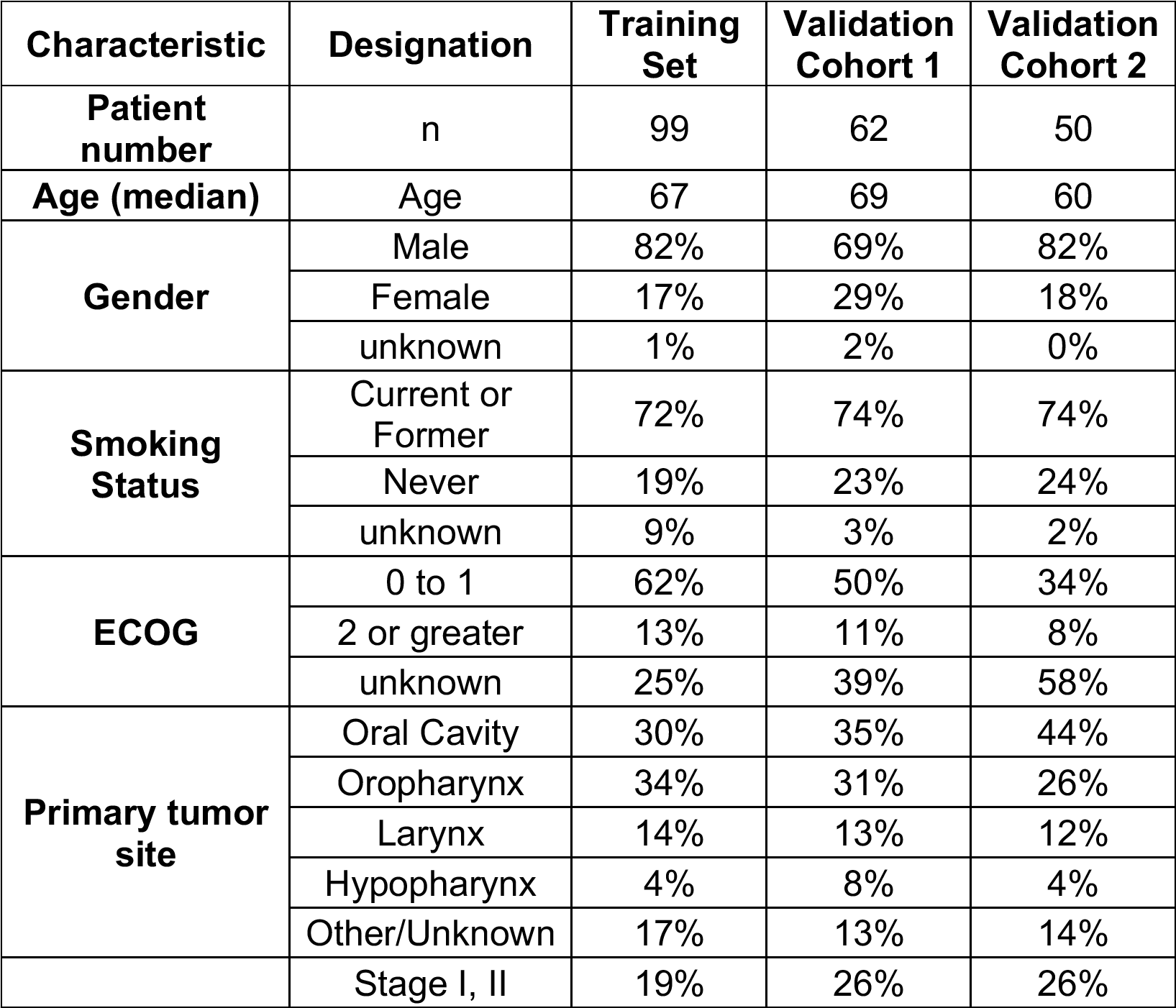
Patient Data.

Validation samples were processed with OncoPrism-HNSCC, generating an OncoPrism Score and resulting OncoPrism Group for each patient sample. Analytical variation was low, with highly repeatable results for replicate samples (manuscript in preparation). The primary endpoint of this study was disease control, so performance was evaluated using the DCR for each OncoPrism Group, with an expected trend from lower DCR in the Low Group to higher DCR in the High Group.

### Validation Cohort 1: monotherapy patients

Specimens from 62 patients treated with anti-PD-1 monotherapy (Cohort 1) were scored and categorized into the Low, Medium, or High Group based on their score and the pre-established thresholds (Figure 2A). The groups roughly mirrored the expected population distribution, with 27% of patients in the Low Group, 32% of patients in the Medium Group, and 40% of patients in the High Group (Table 2).

**Table 2:**
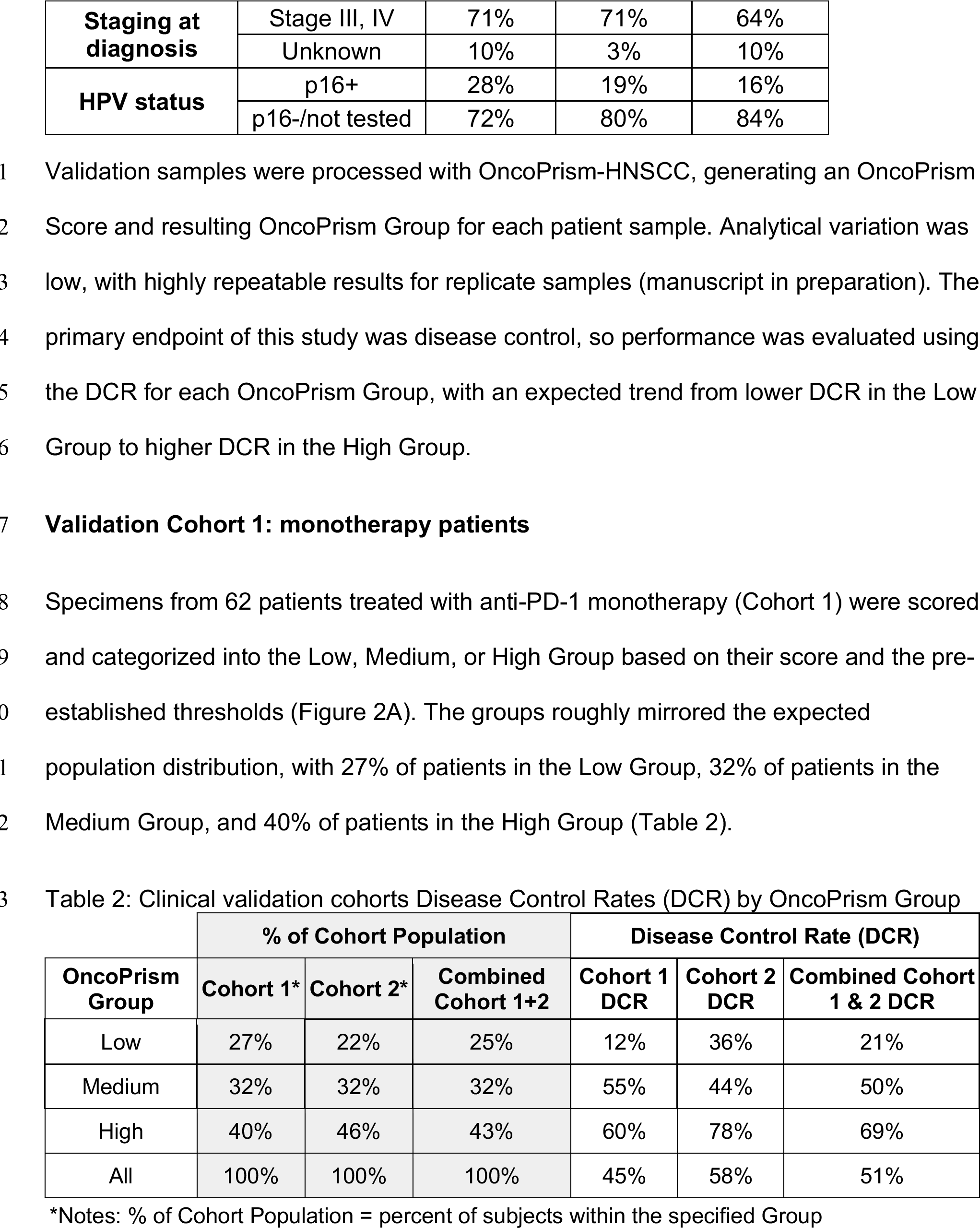
Clinical validation cohorts Disease Control Rates (DCR) by OncoPrism Group.

In Cohort 1 the DCR increases from OncoPrism Low to Medium to High Groups (Table 2; Figure 2B; Cochran-Armitage trend p=0.004). In addition to higher DCR, patients in the High Group also had significantly longer progression-free survival (PFS; log rank test, p=0.015). Median PFS was 2.6 months for the Low Group, 4.2 months for the Medium Group, and 9.8 months for the High Group (Figure 2C). Table 3 shows key performance metrics when comparing the Low Group vs. the High Group, the two actionable OncoPrism Groups. OncoPrism Medium patients were excluded from these calculations as an indeterminate result (see Methods). OncoPrism-HNSCC predicted disease control with high accuracy (0.71), high sensitivity (0.88), specificity (0.60), positive predictive value (PPV [0.60]), and negative predictive value (NPV [0.88]).

**Table 3:** Performance metrics for validation cohorts.

| Cohort | Accuracy | Sensitivity | Specificity | PPV* | NPV* |
| --- | --- | --- | --- | --- | --- |
| Cohort 1: monotherapy<br>(95% Confidence Interval)<br>[Low vs High] | 71%<br>(58-86%) | 88%<br>(74-100%) | 60%<br>(39-83%) | 60%<br>(39-81%) | 88%<br>(75-100%) |
| Cohort 2: chemo-immunotherapy<br>(95% Confidence Interval)<br>[Low vs High] | 74%<br>(62-88%) | 82%<br>(65-96%) | 58%<br>(31-89%) | 78%<br>(62-95%) | 64%<br>(33-89%) |

\*Notes: PPV = Positive Predictive Power; NPV = Negative Predictive Power

### Validation Cohort 2: chemo-immunotherapy patients

To test the ability of OncoPrism-HNSCC to predict disease control in these chemo- immunotherapy-treated patients, performance of OncoPrism-HNSCC was evaluated in specimens from 50 patients treated with anti-PD-1 in combination with chemotherapy (Cohort 2). The groups roughly mirrored the expected population distribution, with 22% of patients in the Low Group, 32% of patients in the Medium Group, and 46% of patients in the High Group (Table 2; Figure 2D). As expected, the overall DCR for this cohort was higher than the monotherapy cohort, likely due to the additional effect of chemotherapy on outcome (58% vs. 45%; Table 2). As with Cohort 1, the DCR increases from OncoPrism Low to Medium to High Groups (Table 2; Figure 2E; Cochran-Armitage trend, p=0.01). This trend corresponded with significantly longer PFS for patients in the High Group (log rank test, p=0.037). Median PFS was 3.0 months for the Low Group, 3.4 months for the Medium Group, and 16.3 months for the High Group in this cohort (Figure 2F). One patient from the High Group was excluded due to an unknown date of progression (n=49). OncoPrism-HNSCC predicted disease control with high accuracy (0.74), high sensitivity (0.82), specificity (0.58), PPV (0.78), and NPV (0.64) when treating the OncoPrism High Group as the predicted positive class and the OncoPrism Low Group as the predicted negative class (with the OncoPrism Medium Group excluded as an indeterminate result; Table 3).

### OncoPrism-HNSCC is not predictive in non-ICI datasets

Our data shows that the OncoPrism Group assignment is correlated with DCR in patients treated with ICIs. To explore whether OncoPrism Group is predictive of disease control in response to ICI or simply prognostic of outcome regardless of therapy, we used the underlying OncoPrism-HNSCC model on four publicly available datasets of HNSCC patients who were not treated with ICI [16,17,21,22]. The OncoPrism-HSNCC biomarker was not significantly correlated with overall survival in any of the non-ICI datasets (Table S4), suggesting that it is not an overall prognostic biomarker *per se* and is consistent with the idea that OncoPrism-HSNCC is predictive of ICI disease control specifically.

### OncoPrism-HNSCC outperforms the existing biomarkers PD-L1 CPS and TMB

Currently, the biomarkers most frequently used to predict response to ICI in RM-HNSCC patients are PD-L1 CPS and, less commonly, Tumor Mutational Burden (TMB). Using our two validation cohorts, we compared the performance of OncoPrism-HNSCC to PD-L1 CPS and TMB. First, we compared PD-L1 CPS to OncoPrism-HNSCC at all possible thresholds for each biomarker using Receiver Operating Characteristic (ROC) curves. For monotherapy-treated Cohort 1, the area under the curve (AUC) for OncoPrism-HNSCC was 0.73, compared to 0.62 for PD-L1 CPS (Figure 3A). Likewise, for the chemo-immunotherapy-treated Cohort 2, the OncoPrism-HNSCC AUC was 0.73 compared to 0.61 for PD-L1 CPS (Figure 3B).

**Figure 3.**
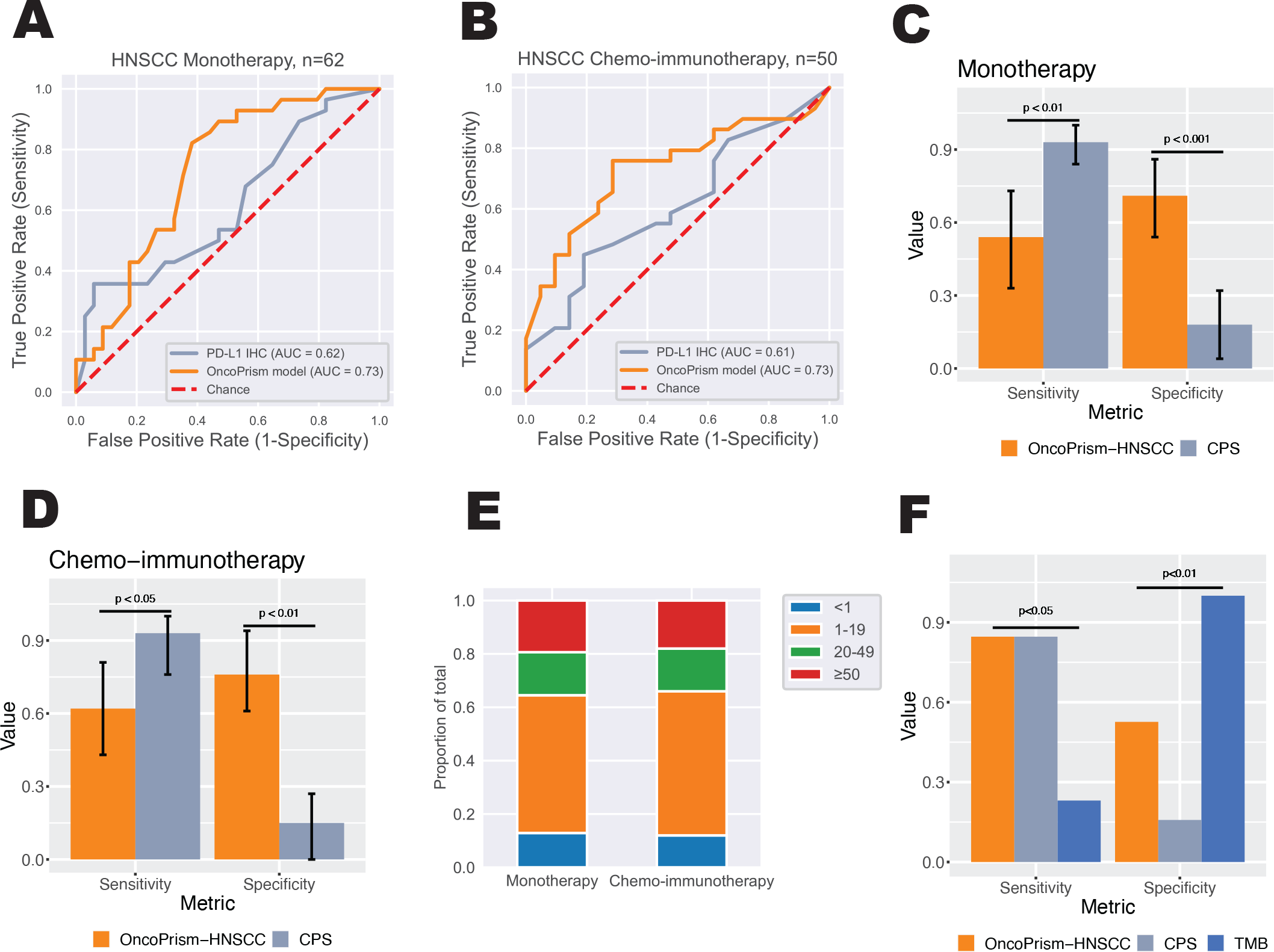
OncoPrism-HNSCC outperforms existing assays PD-L1 CPS and TMB. Receiver Operating Characteristic (ROC) curves are shown for the monotherapy **(A)** and chemo-immunotherapy **(B)** cohorts. OncoPrism-HNSCC (orange) has a higher area under the curve (AUC) than PD-L1 CPS (grey) in both cohorts. In monotherapy **(C)** and chemo-immunotherapy **(D)** cohorts, OncoPrism-HNSCC (orange) has high sensitivity and specificity, while PD-L1 CPS (grey) has high sensitivity but low specificity. The distribution of CPS is similar in each validation cohort **(E)**. **(F)** Sensitivity and specificity for OncoPrism-HNSCC (orange), PD-L1 CPS (grey), and TMB (blue) in 32 patients from the OncoPrism High and Low Groups. Error bars represent 95% Confidence Intervals.

Examining ROC curves is useful since each test has its own thresholds for dividing groups. However, it is also important to compare performance using the commonly used thresholds for each test. For OncoPrism-HNSCC, these thresholds are the divisions between the Low, Medium, and High OncoPrism Groups. For PD-L1 CPS, we categorized patients as PD-L1 Negative (CPS<1) or PD-L1 Positive (CPS≥1), the threshold recommended by ASCO guidelines[23]. In Cohort 1, the DCR for PD-L1 Negative patients was 25% compared to 48% for PD-L1 Positive patients. In Cohort 2, PD-L1 Negative patients had a DCR of 50% compared to 59% for PD-L1 Positive. PD- L1 status was not correlated with PFS in either cohort (Figure S1).

To compare sensitivity and specificity between OncoPrism-HNSCC and PD-L1 in the same population, the OncoPrism High Group was designated as predicted disease control (predicted positive class), while the OncoPrism Medium and Low Groups were designated as predicted disease progression (predicted negative class). This strategy differs from the metrics shown in Table 3, where the Medium Group was excluded as indeterminate in order to match the intended use of the test, but it allows calculation of metrics in the same patient populations for direct comparison. In the monotherapy cohort (Figure 3C), OncoPrism-HNSCC had a sensitivity of 0.54 and a specificity of 0.71. The lower sensitivity compared to Table 3 is due to the inclusion of the Medium Group. Using CPS≥1 to define predicted disease control, PD-L1 CPS had a sensitivity of 0.93 in this cohort. However, the OncoPrism-HNSCC specificity of 0.71 is significantly higher than the CPS specificity of 0.18 (McNemar’s test, p<0.001). Likewise, in the chemo-immunotherapy cohort (Figure 3D), OncoPrism-HNSCC had a sensitivity of 0.62, compared to 0.90 for PD-L1 CPS (McNemar’s test, p<0.05). Again, the OncoPrism-HNSCC specificity of 0.76 is significantly higher than the PD-L1 CPS specificity of 0.14 (McNemar’s test, p<0.001; Figures 3C-D & Table S5). Reflecting the relative sensitivities and specificities of each test, OncoPrism-HNSCC had more false negatives while PD-L1 CPS had more false positives, although 71% of the OncoPrism- HNSCC false negatives were in the Medium Group and would typically be treated as an indeterminate result (Table S6). Interestingly, the proportions of patients in each CPS category was very similar between Cohort 1 and Cohort 2, suggesting that the CPS result was not influencing the treatment of the patients and skewing the datasets (Figure 3E).

Because PD-L1 CPS has low specificity, clinicians may have limited confidence that a PD-L1 Positive patient will indeed benefit from ICI. To test whether OncoPrism-HNSCC can predict PD-L1 Positive patients who have clinical benefit from ICI, we evaluated PFS for each OncoPrism Group in PD-L1 Positive patients only (combined monotherapy and chemo-immunotherapy cohorts). OncoPrism High patients had significantly longer PFS than OncoPrism Medium or Low patients (log rank test, p<0.001; Figure S2).

TMB is less commonly used to guide treatment decisions in RM-HNSCC, but is recommended in some tumor types and when PD-L1 CPS is not available[23]. To compare the performance of OncoPrism-HNSCC to TMB, we evaluated TMB status for samples from the monotherapy-treated cohort (Cohort 1). Specifically, all OncoPrism High or Low samples with sufficient material were evaluated (32 samples in total). We evaluated samples in the High and Low Groups as these two categories are the most likely OncoPrism-HNSCC test results to influence a clinical decision. TMB of at least 20 mut/Mb was classified as TMB High, while less than 20 mut/Mb was considered TMB Low (see Methods). Overall, OncoPrism-HNSCC had a sensitivity of 0.85 and a specificity of 0.53 in this group, compared to a sensitivity of 0.23 and a specificity of 1 for TMB (Figure 3F). The sensitivity and specificity for CPS in this group is also shown for reference. While TMB had significantly higher specificity than OncoPrism-HNSCC (McNemar’s test, p=0.008), it only identified three patients with disease control (Table S7). OncoPrism-HNSCC had significantly higher sensitivity than TMB (McNemar’s test, p=0.027).

## DISCUSSION

OncoPrism-HNSCC significantly predicts disease control and PFS in response to anti-PD-1 (ICI) therapy in pre-treatment RM-HNSCC patients. Importantly, the test was validated in two separate cohorts using patient samples from 17 clinical academic and community sites from across the United States, which allowed us to account for test performance across a variety of possible pre-analytic sample processing conditions.

The multi-dimensional biomarker underlying OncoPrism-HNSCC was built using the careful evaluation of a previously published study of cell composition, cell-state, and immune modulatory genes in the tumor microenvironment [14]. Both validation cohorts (Cohort 1: monotherapy and Cohort 2: chemo-immunotherapy) had similar results, with a significant correlation of OncoPrism Group classification with DCR and PFS, as well as high accuracy, sensitivity, specificity, PPV, and NPV. The OncoPrism-HNSCC model was not predictive in patients treated with non-ICI therapies, suggesting that the biomarker is predictive rather than prognostic (Table S4). These results also suggest that the predictive nature of the biomarker may be specific to the ICI component in chemo-immunotherapy-treated Cohort 2.

There was no distinct difference observed in PD-L1 CPS status of those patients prescribed monotherapy (Cohort 1) vs. chemo-immunotherapy (Cohort 2) (Figure 3E), suggesting that PD-L1 score was not driving treatment decision between monotherapy and chemo-immunotherapy in this study. Limitations of PD-L1 for guiding treatment have been previously published[24,25].

The intended use of OncoPrism-HNSCC is to aid clinicians in choosing whether to treat with anti-PD-1 as a single agent, anti-PD-1 in combination with chemotherapy, or alternative treatment options. While patients treated with chemo-immunotherapy have higher DCR than monotherapy-treated patients, chemo-immunotherapy is also associated with higher toxicity[4,9,26]. OncoPrism-HNSCC improves upon other assays to inform these treatment decisions, with the potential to limit unnecessary treatment-associated toxicities and improve patient outcomes. Currently, PD-L1 CPS is the most common biomarker used to guide such decisions. Unfortunately, PD-L1 CPS has high sensitivity but low specificity for predicting disease control (Fig 3C-D and Table S5).

This low specificity means that many patients with high CPS do not clinically benefit from ICI, and clinicians are reluctant to use the CPS to exclude patients from more aggressive treatment options like chemo-immunotherapy. OncoPrism-HNSCC has significantly higher specificity than PD-L1. In addition, OncoPrism Groups stratify patients by PFS among all patients (Figure 2C&F) and in PD-L1 Positive patients alone (Figure S2). Together, these results suggest that ICI therapy should be prioritized for OncoPrism High patients (Figure 4). Given the low DCR and PFS in the OncoPrism Low Group, clinicians should consider a non-ICI treatment and/or available clinical trials for OncoPrism-HNSCC Low patients regardless of PD-L1 status. While PD-L1 CPS had significantly higher sensitivity than OncoPrism-HNSCC when categorizing OncoPrism Low and Medium patients as predicted progressors (Figure 3C-D), OncoPrism-HNSCC had similar sensitivity to PD-L1 when comparing OncoPrism Low to High (see Methods; Table 3 and Table S5). In contrast to PD-L1, TMB had high specificity but low sensitivity (Figure 3F). As a result, ICI treatment should be prioritized for TMB High patients, but OncoPrism-HNSCC results appear to have superior prediction over TMB Low results.

**Figure 4.**
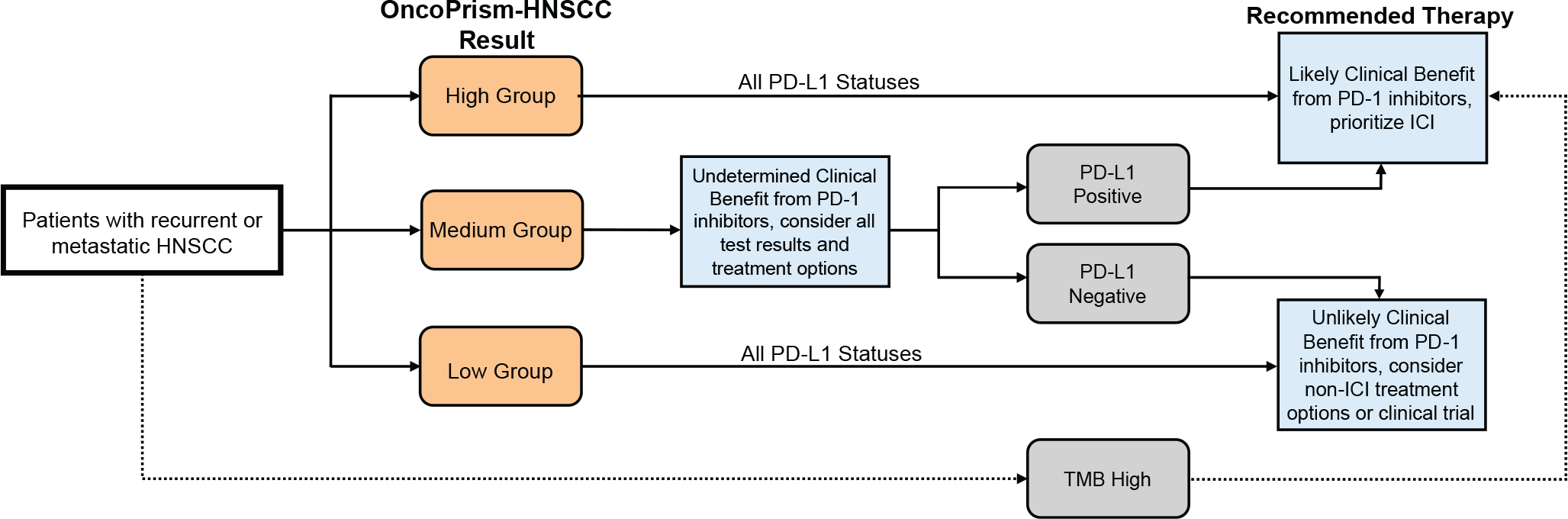
Immune checkpoint inhibitor decision tree based on test results. Patients with recurrent or metastatic HNSCC tested with OncoPrism-HNSCC are categorized into the OncoPrism Low, Medium, or High Group. Because OncoPrism-HNSCC has high specificity relative to PD-L1 CPS and OncoPrism High patients have longer PFS regardless of PD-L1 status, OncoPrism High patients should typically be treated with ICI regardless of PD-L1 status. OncoPrism Low patients have low ICI DCR and are not good candidates for ICI even if they are PD-L1 Positive. Patients in the OncoPrism Medium Group do not have a definitive treatment path; all test results and treatment options should be considered. Typically, ICI should be favored for OncoPrism Medium patients who are PD-L1 Positive while non-ICI or clinical trial options should be considered for PD-L1 Negative patients. TMB testing is not recommended for most HNSCC patients. However, if TMB testing is performed, ICIs should be prioritized for TMB High patients given the high observed specificity of TMB. Only 9% of patients in our study were TMB High. Due to the low sensitivity of TMB, a TMB Low result should not be strongly considered in treatment decisions.

Figure S3 provides a summary of treatment recommendations based on test results.

This real-world observational validation study has several limitations. These limitations include patients with long time between test biopsy and treatment, the inclusion of patients who had intervening treatments between the biopsy and the ICI, and instances of incomplete data within the patient’s clinical record. The inclusion criteria in this study balanced recruiting real-world, well-controlled patient cohorts reflecting the intended treatment scenario with maximizing the study size. Ongoing and future studies aim to refine the model and study additional endpoints such as overall survival with additional patient cohorts. In addition, future work will refine the thresholds for each OncoPrism Group to maximize the number of patients with actionable results without sacrificing the sensitivity or specificity of the test. The Medium Group is considered an indeterminate result due to the variation seen in Medium Group DCR and PFS across the training cohort and validation cohorts (data not shown and Fig 2B-C & E-F). As a result, currently the Medium Group result does not provide clear clinical guidance to physicians and patients. In the validation cohorts, 32% of patients fell in the Medium Group, meaning 68% of patients would have received treatment-guiding information. Because OncoPrism-HNSCC balances high sensitivity and high specificity for predicting disease control, it is clinically useful and has the potential to aid treatment decisions of more patients than existing tests.

Although there is no ICI predictive biomarker with perfect sensitivity and specificity, OncoPrism-HNSCC addresses significant shortcomings of PD-L1 and TMB in the RM-HNSCC population through a balance of sensitivity and specificity, enabling clinicians to identify patients most likely to benefit from immunotherapy. OncoPrism-HNSCC exhibits clinical validity across a diverse patient population and holds promise to guide treatment decisions and improve patient outcomes.

## DECLARATIONS

### Ethics approval and consent to participate

The study protocol was approved through WCG or Advarra IRB and/or at the local site level IRB. Independent data monitoring was conducted by the study clinical research organization Curebase, Inc (San Francisco, CA). All patients provided signed, informed consent to participate, or consent was waived for deceased patients according to the approved study protocol.

### Consent for publication

Not applicable

### Availability of data and material

The data underlying this study, including anonymized patient-level OncoPrism-HNSCC, PD-L1, and TMB measurements, are available for non-commercial use from the corresponding author upon reasonable request. Materials and analytical code are not available.

### Authors’ contributions

Conception and design: KCF, JE, JH, RLW, MMP, DNM, JIG, EJD

Recruitment of patients and acquisition of samples and data: MMP, HM, VV, LS, ID, OKM, KW, JLW, GA, AWP, JS, BB, XS, SEK, TDS, WLA, KAP, JL, DNM, JIG, ADC, EEWC, DRA,

Execution of the research: KCF, JE, JH, RLW, MMP, WHW, EJD Analysis and interpretation of data: KCF, JE, JH, EJD

Drafting of the manuscript: KCF, JE, JIG

## Data Availability

The data underlying this study, including anonymized patient-level OncoPrism-HNSCC, PD-L1, and TMB measurements, are available for non-commercial use from the corresponding author upon reasonable request.

## ACKNOWLEDGMENTS

We would like to thank the patients who participated in this study. In addition, we thank all members of the PREDAPT study consortium. In particular, we thank the following people for their invaluable contributions: Courtney Alexander, Rachel Allen, Nowsheen Azeemuddin, Rajesh Bande, Brandon Barker, Savanna Biedermann, Darcie A. Cruz, Karl D’Silva, Paula Datri, Deirdre L. Dillon, Julie A. Faust, Eileen M. Georgi, Paul Gonzales, Carrie Grandidier, Leah A. Guilford-Elenes, Ann Hargrove, Peter Jiang, Paraic A. Kenny, Cheryl L. Krantz, Mukesh Kumar, Natalie K. Kyek, Steven Lai, Amber N. Larimer, Jenna R. Larry, Robert C. Matulonis, Elyssa Navarro, Uyen Nguyen, Suresh Nukala, Lucyna Olechny, Meley L. Pine, Breanna N. Richardson, Julie Roache, Destiny Robinson, Steven Saccaro, Alaina B. Sandoz, Kelly Shrimpton, Julie Skarsvog, Peggy Smith, Scott A. Sonnier, Galen A. Steinhoff, Jennifer Taylor, Alierykel Williams, Erin L. Wilson, and Anna Young.

---

Figure S1. PD-L1 does not predict progression-free survival (PFS). Patients were divided into PD-L1 Positive (CPS≥1) or Negative (CPS<1) groups within each validation cohort. PD-L1 status was not significantly correlated with PFS in the monotherapy (p=0.71) **(A)** or chemo-immunotherapy (p=0.28) **(F)** validation cohorts. P-values for PFS were calculated using log rank methods.

Figure S2. OncoPrism-HNSCC predicts progression-free survival (PFS) in PD-L1 Positive patients. All PD-L1 Positive patients from both Cohort 1(monotherapy) and Cohort 2 (chemo-immunotherapy) were grouped according to their OncoPrism-HNSCC result. OncoPrism High Group is significantly associated with longer PFS (p<0.001, log rank methods).

Figure S3. Treatment prioritization for discordant test results. The recommended treatment for concordant (grey) and discordant (white) test results is shown for OncoPrism-HNSCC compared to PD-L1 CPS (**A**) or TMB (**B**). The percent shown is the percentage of the study population in each category. The most common discordant test results are outlined for PD-L1 (red) and TMB (blue). OncoPrism Medium samples were considered indeterminate and are not included. For comparison with PD-L1, n=75. For comparison with TMB, n=32.

