## Supplemental Material for "Multicenter validation of an assay to predict anti-PD-1 disease control in patients with recurrent or metastatic Head and Neck Squamous Cell Carcinoma: The PREDAPT Study"

Table S1: Excluded Patient Samples

| | |
|---|---|
| None (included in study) | 211 |
| Unacceptable Treatment Regimen | 39 |
| Excessive Time to Treatment | 18 |
| Unacceptable Specimen Site | 17 |
| Incomplete Data | 14 |
| Insufficient Material | 10 |
| Other Ineligible* | 18 |
| Tumor cellularity | 48 |
| Fail RNA QC | 13 |
| Fail Library QC | 10 |
| Fail Analytical QC | 15 |
| Withdrawn | 2 |
| **Total** | **415** |

*Note:*
*Includes excluded diagnoses, inconsistent data, site sending wrong material, and other violations of inclusion/exclusion criteria

### Table S2. HUGO gene designations of input genes for model development

| | | | | | | | |
|---|---|---|---|---|---|---|---|
| C2 | PTPRC | HLA-DRA | C1QB | CD3E | FCER1G | C3AR1 | CCL4 |
| DOCK8 | LILRB4 | CCR5 | CCL3L3 | CXCR3 | GZMH | GPR65 | CD80 |
| STAT2 | DRAM1 | IL21R | TNFAIP3 | PSMB9 | IL15 | GBP5 | CD300LF |
| RNF19B | ETV7 | CD96 | CXCL11 | TMEM140 | PLAUR | ISG20 | IFNG |
| SLC15A3 | GPR171 | APOL3 | RIPK2 | LILRB2 | SLA2 | ICAM1 | SPI1 |
| IFIT2 | SIRPG | CCL5 | CD69 | IL15RA | TRANK1 | CD74 | DOK2 |
| IFI30 | TAP2 | IL2RB | TRIM21 | HLA-B | WARS | GBP2 | STAT4 |
| CXCL10 | PARP14 | CD163 | SP100 | SAMSN1 | PDCD1 | CD2 | CD300A |
| JAK2 | FPR3 | UHRF2 | NLRC5 | UBA7 | CSF1 | LAG3 | RANBP6 |
| HAPLN3 | SH2D1A | DNAJA1 | ZBP1 | BTN3A1 | IL18BP | CTSL | |
| PLGRKT | SRGN | BATF2 | SIGLEC10 | CXCR6 | PDCD1LG2 | CD3G | |
| CDC37L1 | UBE2L6 | CD274 | IDO1 | DDX58 | SAMHD1 | GPR84 | |
| EPSTI1 | TIGIT | IFIH1 | BTN3A3 | PPP1R18 | ITGAL | CIITA | |
| HLA-F | FCGR1A | TNFRSF9 | GZMA | GBP1 | CCL8 | C1QC | |
| ITK | STAT1 | SP110 | TRIM22 | FGL2 | STX11 | TAP1 | |
| GNLY | SLAMF8 | GZMB | HLA-DRB1 | NKG7 | LAP3 | TNFSF13B | |
| LCP2 | IL2RA | FCGR3A | PRF1 | GBP3 | CTLA4 | IRF1 | |
| HAVCR2 | APOL6 | IFIT3 | IL12RB1 | MYO7A | CD86 | ICOS | |
| FASLG | GBP4 | TLR8 | CCR1 | STK10 | CXCL9 | PLA2G7 | |
| RNF213 | CD8A | FCGR2A | PTPN22 | B2M | RIC1 | SAMD9L | |

### Table S3. Inclusion/exclusion criteria

| | |
|---|---|
| Inclusion criteria: | |
| 1. | Subject must have been diagnosed with recurrent or metastatic head and neck squamous cell carcinoma (RM-HNSCC). |
| 2. | Subject must have received, or been scheduled to receive, at least one dose of anti-PD-1 immunotherapy for treatment of their cancer. |
| 3. | Subject must have received, or been scheduled to receive, anti-PD-1 treatment on one of the following regimes:<br>A. Pembrolizumab or nivolumab as a single agent (monotherapy)<br>B. Pembrolizumab or nivolumab in combination with platinum chemotherapy, with or without fluorouracil or a taxane (chemo-immunotherapy) |
| 4. | Subject must have had, or will have had, a tumor biopsy prior to treatment with anti-PD-1 immunotherapy (specimen is considered pre-treatment). |
| 5. | Clinician must have evaluated, or will have evaluated, tumor response to anti-PD-1 immunotherapy using (1) imaging data performed to assess response to anti-PD-1 treatment and/or (2) clinical assessment. Imaging data must include both (a) pre-anti-PD-1-treatment imaging and (b) imaging performed after the start of anti-PD-1 treatment). |
| 6. | Subject is willing to provide informed consent per IRB-approved protocol. |
| 7. | Subject must have sufficient tissue available to fulfill the specimen requirements of the study. |
| 8. | Subject is 18 years of age or older. |
| Exclusion criteria: | |
| 1. | Subjects who did not have head and neck squamous cell carcinoma (other histologies). |
| 2. | Subjects who only received anti-PD-1 immunotherapy in the newly diagnosed, curative setting. |
| 3. | Subjects who received anti-PD-1 immunotherapy in combination with a treatment other than platinum chemotherapy (with or without fluorouracil or a taxane). |
| 4. | Subjects whose response to anti-PD-1 immunotherapy is not available either as (a) clinician’s evaluation of that response or (b) imaging data that was performed to assess response to anti-PD-1 immunotherapy. |

**Table S4.** OncoPrism-HNSCC model is not predictive in non-ICI datasets

| dataset | Number of samples | OncoPrism-HNSCC model |
|---|---|---|
| TCGA-HNSC | 500 | 0.32 |
| GSE65858 | 290 | 0.66 |
| GSE40774 | 90 | 0.56 |
| GSE41613 | 97 | 0.71 |

*Notes: the p-values represent p-values for log rank test done on a Cox proportional hazards model.

**Table S5.** PD-L1 performance metrics

| Cohort | Comparison | Accuracy | PPV* | NPV* | Sensitivity | Specificity |
|---|---|---|---|---|---|---|
| Cohort 1 (monotherapy) | CPS<1 vs CPS≥1 | 52% | 48% | 75% | 93% | 18% |
| Cohort 1 (monotherapy) | CPS<20 vs CPS≥20 | 58% | 55% | 60% | 43% | 71% |
| Cohort 2 (chemo-immunotherapy) | CPS<1 vs CPS≥1 | 60% | 60% | 60% | 93% | 15% |
| Cohort 2 (chemo-immunotherapy) | CPS<20 vs CPS≥20 | 60% | 76% | 52% | 46% | 80% |

*Notes: PPV=positive predictive value; NPV=negative predictive value

Table S6: Comparison of OncoPrism and PD-L1 CPS

| | PD-L1 TP | PD-L1 FN | PD-L1 TN | PD-L1 FP |
|---|---|---|---|---|
| OncoPrism TP | 32 | 1 | - | - |
| OncoPrism FN | 21 | 3 | - | - |
| OncoPrism TN | - | - | 8 | 32 |
| OncoPrism FP | - | - | 1 | 14 |

*Abbreviations:*

TP, True Positive, correctly predicted disease control;

FN, False Negative, disease control incorrectly predicted as no disease control;

TN, True Negative, correctly predicted no disease control;

FP, False Positive, no disease control incorrectly predicted as disease control

Table S7: Comparison of OncoPrism and TMB

| | TMB TP TP | TMB FN | TMB TN | TMB FP |
|---|---|---|---|---|
| OncoPrism TP | 2 | 9 | - | - |
| OncoPrism FN | 1 | 1 | - | - |
| OncoPrism TN | - | - | 10 | 0 |
| OncoPrism FP | - | - | 9 | 0 |

*Abbreviations:*
TP, True Positive, correctly predicted disease control;
FN, False Negative, disease control incorrectly predicted as no disease control;
TN, True Negative, correctly predicted no disease control;
FP, False Positive, no disease control incorrectly predicted as disease control

### Figure S1: PD-L1 does not predict PFS

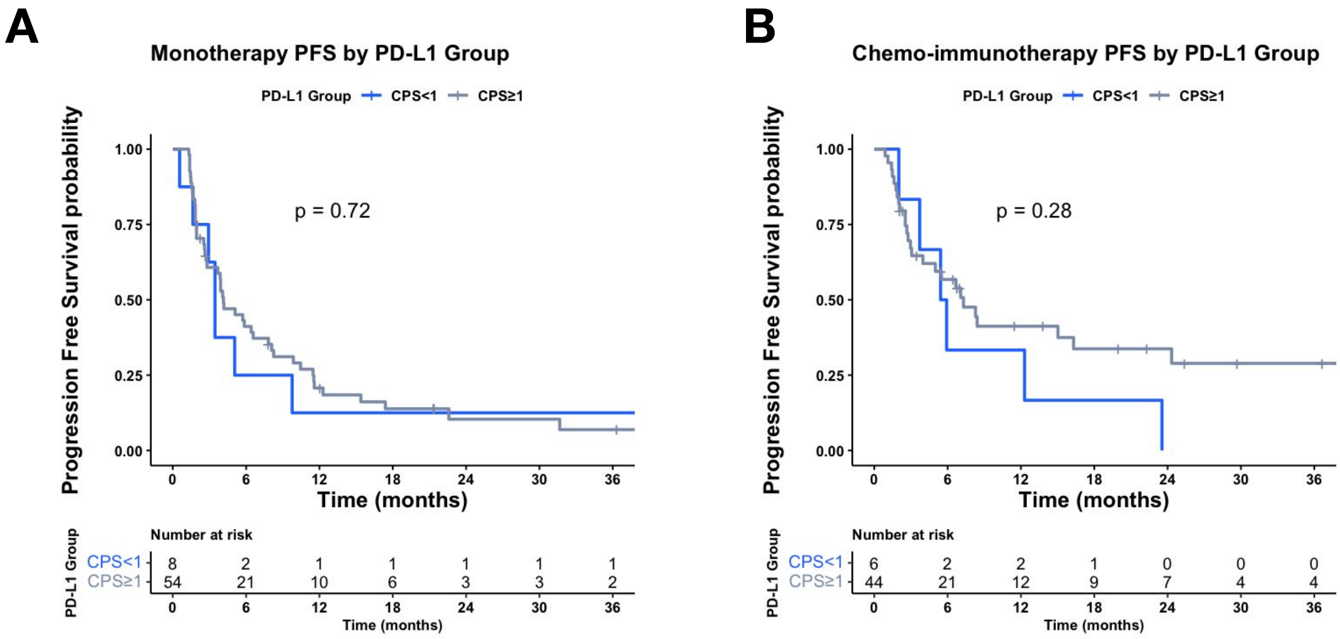

**Figure S2: OncoPrism-HNSCC predicts PFS in PD-L1 positive patients**

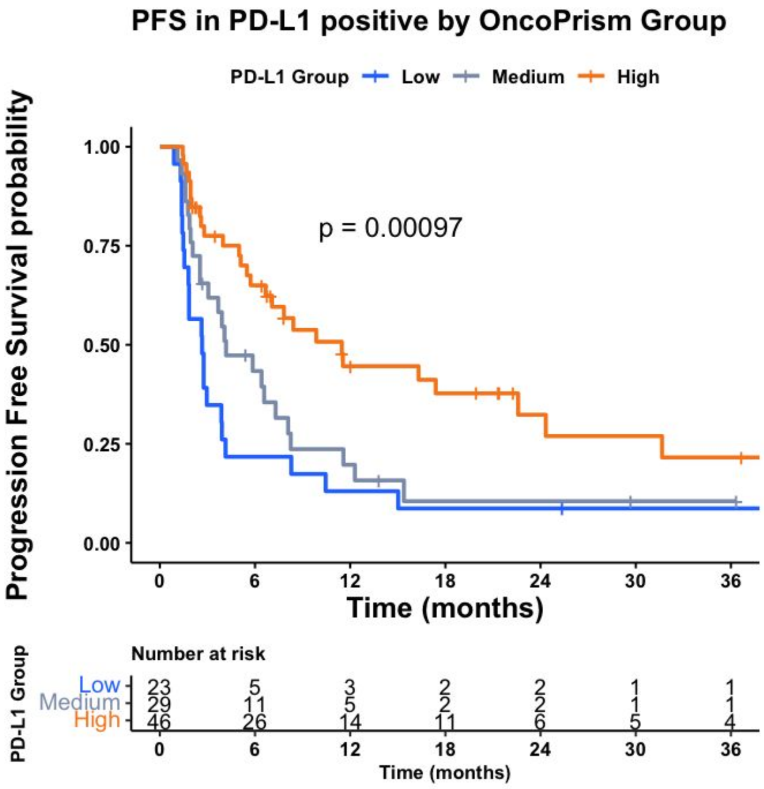

**Figure S3.** Discordant PD-L1/TMB Rate and Treatment Prioritization

**A**

PD-L1 Discordance Treatment Prioritization

| | PD-L1 (-) | PD-L1 (+) |
|---|---|---|
| OncoPrism-HNSCC Low | **non-ICI** (7% of study) | **non-ICI** (31% of study) |
| OncoPrism-HNSCC High | **ICI** (3% of study) | **ICI** (60% of study) |

**B**

TMB Discordance Treatment Prioritization

| | TMB-Low | TMB-High |
|---|---|---|
| OncoPrism-HNSCC Low | **non-ICI** (34% of study) | **non-ICI** (3% of study) |
| OncoPrism-HNSCC High | **ICI** (56% of study) | **ICI** (6% of study) |
